# Characterizing breast cancer response to neoadjuvant therapy based on biophysical modeling and multiparametric magnetic resonance imaging data

**DOI:** 10.1101/2023.11.28.23299112

**Authors:** Haley J. Bowers, John A. Guthrie, Alaura Krukoski, Alexandra Thomas, Jared A. Weis

## Abstract

Personalized medicine efforts are focused on identifying biomarkers to guide individualizing neoadjuvant therapy regimens. In this work, we aim to validate a previously developed image data-driven mathematical modeling approach for dynamic characterization of breast cancer response to neoadjuvant therapy using a large, multi-site cohort. We retrospectively analyzed patients enrolled in the BMMR2 ACRIN 6698 subset at 10 institutions. Patients enrolled received four MRI examinations during neoadjuvant therapy with acquisitions at baseline (T_0_), 3-weeks/early-treatment (T_1_), 12-weeks/mid-treatment (T_2_), and completion of therapy prior to surgery (T_3_). A biophysical mathematical model of tumor growth is used extract metrics to characterize the dynamics of treatment response. Using predicted response at therapy conclusion and histogram summary metrics to quantify estimated tumor proliferation maps, we found univariate model-based metrics able to predict pathological response, with area under the receiver operating characteristic curve (AUC) ranging from 0.58 and 0.69 analyzing between T_0_ and T_1_, and AUCs ranging from 0.72-0.76 analyzing between T_0_ and T_2_. For hormone receptor (HR)-negative, human epidermal growth factor receptor 2 (HER2)-positive breast cancer patients our model-based metrics achieved an AUC of 0.9 analyzing between T_0_ and T_1_ and AUC of 1.0 analyzing between T_0_ and T_2_. This data shows the significant promise in developing these imaging-based biophysical mathematical modeling methods of dynamic characterization into a clinical decision support tool for individualizing treatment regimens based on patient-specific response.

## Introduction

The goal of personalized medicine is to revolutionize healthcare by focusing on matching the most accurate and efficient treatment regimen to the uniqueness of an individual patient and their cancer. The majority of efforts focus on utilizing genomic sequence information to identify diagnostic and prognostic features for risk assessments^1^. Molecular approaches to cancer therapy have advanced cancer care by guiding therapy selection based on gene/protein expression but ignore other important patient-specific phenotypic properties that influence response to therapy including tumor microenvironment and patient specific factors such as dose exposure and dose intensity. Patient-specific tumor characteristics, such as spatial and temporal phenotypic variability^2^, encode additional insights for individualizing treatment regimens and response to therapy. Evaluation of such properties could support new hypotheses regarding optimal dosing and therapeutic regimen scheduling. Optimal dosing and regimens have the potential to guide therapy de-escalation and escalation strategies to optimize cytotoxic therapies and minimize treatment-related toxicities^3–5^. Unfortunately, current response assessment tools are unable to sufficiently incorporate these patient-specific, phenotypic characteristics, limiting the adaptation of personalized treatment regimens.

Pre-surgical neoadjuvant therapy (NAT) is the standard of care for locally-advanced and high-risk breast cancer. Breast cancer response to NAT is currently assessed using physical examination and/or imaging to assess morphological changes of the tumor. Magnetic resonance imaging (MRI) is often used for assessment pre-NAT to determine the extent of disease and at the conclusion of NAT, prior to surgery, to radiologically estimate response for surgical planning. MRI also holds strong potential to enable more quantitative assessment throughout the NAT regimen setting by integrating multiparametric imaging protocols of tumors and their response. Multiparametric imaging protocols are often utilized for improved lesion detection, and typically include T1-weighted contrast enhanced and diffusion weighted (DW) techniques. Dynamic contrast enhanced (DCE) MRI is able to capture the permeability of the blood vessels that supply a lesion by obtaining T1-weighted images prior, during, and after intravascular contrast agent administration^6^. DW-MRI is a functional imaging technique used to estimate tumor cell density by quantifying the random movement of water molecules in tissue, which is restricted in high-density tissue, such as solid tumors. From DW-MRI imaging, the apparent diffusion coefficient (ADC) is calculated as a quantitative measurement of diffusivity. Since diffusion is hindered in solid tumors, studies have reported significant correlations between ADC and tumor cellularity. However, quantitative breast DW-MRI study results vary widely due to differences in acquisition, scanner hardware, and interpreter variability, limiting the translation of ADC as a clinical biomarker^7, 8^. The majority of these radiological assessment methods also utilize region-of-interest (ROI)-based metrics which eliminate spatial information encoded within the images. As solid tumors exhibit a high degree of temporally varying intra-tumor heterogeneity ^9^, there is significant need for new accurate spatiotemporal informative assessment methods to guide implementation of patient-specific adaptive therapeutic regimens.

We^10–12^ and others^13–15^ believe that the integration of imaging and mathematical modeling has the ability support the development of personalized response assessment through patient-specific tumor characterization, allowing for individualized therapy response assessment and regimen tailoring based on fundamental biophysical principles that underlie observed tumor response to therapy. In previous work^16^, we developed a mechanically-coupled reaction-diffusion model for the parameterization of biophysical metrics of response for the prediction of pathological complete response (pCR), an outcome metric that has shown to be predictive of recurrence risk and overall survival in breast cancer patients^10^. The goal of this work is to apply our framework of estimating spatial phenotypic biophysical parameters of growth for the characterization of therapy response to enable predictions of residual tumor burden. In this work, we deploy this framework with data from the Breast Multiparametric MR Imaging for prediction of Neoadjuvant chemotherapy (NAC) Response (BMMR2) Challenge through the National Cancer Institute (NCI) Quantitative Imaging Network (QIN). The goal of the BMMR2 challenge was to identify image-based biomarkers derived from DW-MRI and DCE-MRI for predicting pCR following neoadjuvant therapy (NAT) for invasive breast cancer.

## Materials and Methods

### Study Cohort

A total of 191 subjects were enrolled in the BMMR2 ACRIN 6698 subset at 10 institutions and who underwent NAT for invasive breast cancer were provided. Figure 1 shows the study timeline for enrolled patients. The ACRIN 6698 study acquired multiparametric MRI examinations with high resolution DCE-MR and DW-MR techniques at four time points over the course of NAT: pre-treatment (T_0_), early treatment after 3 cycles of therapy (T_1_), midtreatment/12 week into therapy T_2_), and post-treatment and prior to surgery (T_3_). The data was retrieved from the BMMR2 challenge hosted through NCI QIN and only included MRI studies T_0_, T_1_, and T_2_. Patients were assigned therapy based on tumor molecular subtype according to receptor status with hormone receptor (HR) status (estrogen receptor or progesterone receptor) and human epidermal growth factor 2 (HER2) receptor expression determined from pretreatment core biopsy by immunohistochemistry under the I-SPY 2 TRIAL protocol. Tumor subtypes were categorized as HR-/HER2-(triple negative), HR+/HER2-, HR-/HER2+, and HR+/HER2+. Figure 1 describes the NAT treatment regimen whereby patients received 12 weekly cycles of anthracycline-based therapy (control) or in combination with one of the experimental agents, followed by 4 cycles of Adriamycin/Cyclophosphamide administered every 3 weeks. Patients classified as HER2-positive also received anti-HER2 agents.

**Figure 1:**
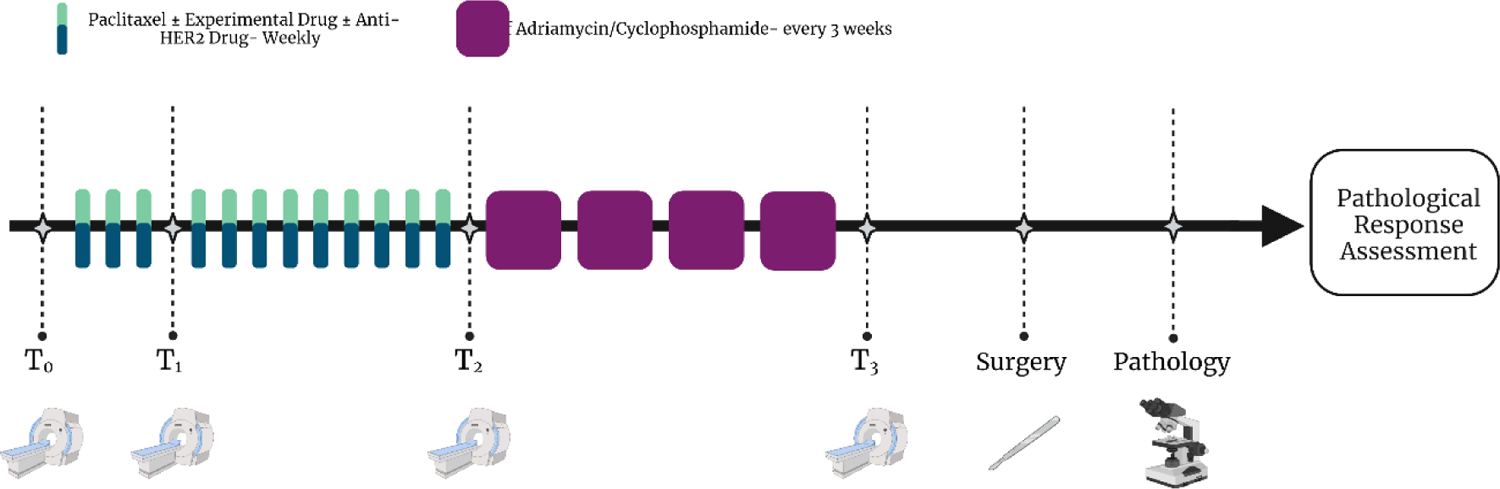
Schematic of the therapeutic and imaging timeline. Patients undergo baseline imaging prior to the start of NAT (T_0_), after three cycles of therapy (T_1_), at the conclusion of the first portion of therapy (T_2_), and at the completion of therapy prior to surgery (T_3_). After surgical resection, pathological analysis is conducted to determine response and/or residual burden of the resected tumor.

### MRI Acquisition

MRI was performed under the original ACRIN 6698 study on 1.5 or 3.0 Tesla scanners. All studies included a localization scan and three bilateral axial acquisition sequences: 1) T2-weighted, 2) Diffusion weighted (DW) (with b= 0, 100, 600, 800 s/mm^2^, 3-directions), and 3) *T1*-weighted dynamic contrast enhanced (DCE) with phase duration between 80 and 100 seconds with at least 8 minutes of continuous post-injection acquisition.

### Imaging Data Analysis

A previously developed mechanically coupled reaction diffusion model was employed to characterize response throughout the course of NAT. While a brief description follows, interested readers can refer to previous studies for a detailed description of the modeling methodology^10–12^. In short, DW-MRI data at each time point were aligned to DCE-MRI data through scanner offset correction and pixel spacing interpolation. DCE-MRI and DW-MRI data were then co-registered across the serial imaging time points with rigid registration using FLIRT^17–19^ followed by non-rigid registration using DRAMMS^20^ using default registration parameters. The tumor midpoint was identified at the midtreatment time point (T_2_) and central-slice images were extracted and used for subsequent analysis. DCE-MRI data was used to create a tumor region-of-interest (ROI) for each time point in the series by first creating a manual segmentation followed by refinement based on voxels that satisfy a functional signal intensity threshold increase of 70% between the pre-contrast and the first post-contrast image^11, 21^. Apparent diffusion coefficient (ADC) maps were derived and provided with the four b-values included in the ACRIN 6698 DW-MRI datasets^22, 23^. Spatiotemporal cellularity, *N*(r, *t*) was estimated using Equation [1] with ADC data for voxels satisfying the DCE-MRI threshold functional criteria of 70% enhancement^21^:

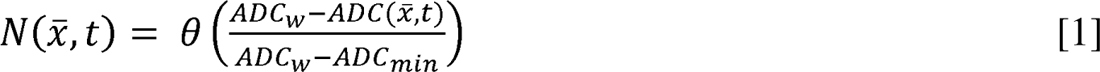

where θ describes the cellular carrying capacity, a geometric constraint on the total number of tumor cells in a voxel, calculated as the ratio of the imaging voxel volume to the assumed tumor cell volume, assuming spherical tumor cells with a packing density of 0.7405 and a nominal tumor cell radius of 10 microns (tumor cell volume of 4189 um^3)^24^.

As described in prior work^11^, the set of coupled, partial differential equations composing the clinical tumor growth model is shown in Equations (2) - (4):

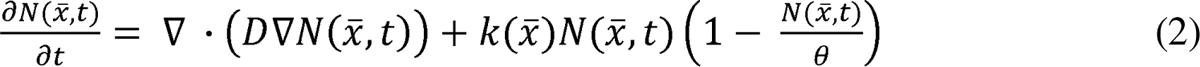

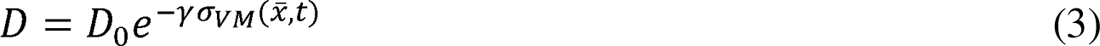

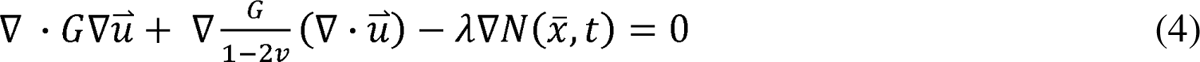

Through estimating the spatiotemporal change in cell number, *N*, we can extract parameter estimates of apparent cell diffusion, *D_0_*, and spatial tumor cell proliferation/death rate, *k*. is Tumor cell diffusion in the absence of external stress, *D_0_*, is estimated through linking *D* to the surrounding tissue mechanics^25^. The system was modeled under the assumption that breast tissue exhibits a linear elastic, isotropic response to mechanical stress. Finite element meshes composed of three-node triangular elements with an average edge length of 1.5 mm were constructed for each patient. To improve computation time for subsequent model-based spatial property reconstruction, the mesh was then clustered into spatial patch regions for property reconstruction using k-means clustering based on Euclidian distance. Regions contain an average of 5 elements per region with average patch area of 3.25 mm^2^. Temporal resolution for the forward problem was assigned at Δ*t* = 1 day.

A schematic of the biophysical parameter characterization framework is shown in Figure 2. Spatial cell number was estimated at early treatment (T_1_) and midtreatment (T_2_) observed imaging time point based on ADC images as described. We assume a piecewise continuous antitumor effect between each observed imaging time point during therapy and characterize phenotypic biophysical parameters between time point combinations: T_01_ (T_0_ and T_1_) and T_02_ (T_0_ and T_2_). This allows for capture of the dynamic changes in parameters between time points as well as evaluation of the importance of intermediate time point imaging acquisitions. The inverse property estimation problem was solved to estimate a region-based spatially varying proliferation rate map and global cell diffusion parameter based on minimizing the error between the model-estimated cellularity and observed cellularity. A quasi-Newton optimization method using L-BFGS was employed to estimate model parameters^26^ with gradients for region-based proliferation calculated using a numerically-efficient adjoint state method^27^ and gradients for diffusion calculated using a forward finite difference method with perturbation of 1%. Following parameter estimation, histograms of the tumor proliferation rate map were created with fixed bin widths of 0.1 days^-1^. We also isolated the growth portion (*k* > 0) of the proliferation histogram for further ‘positive proliferation’ metric extraction. The estimated biophysical parameters were then used in conjunction with the model to project the model forward in time to estimate the residual tumor burden area at the conclusion of NAT, T_3_, (Area_T3,predict_). We also assessed conventional radiologic assessment metrics. We assessed the percent change in metrics between time point combinations: T_01_ (T_0_ and T_1_) and T_02_ (T_0_ and T_2_), for the mean tumor ADC^28^, and functional tumor volume (FTV), a measurement for the tumor volume for lesion regions satisfying the DCE-MRI threshold functional criteria of 70% enhancement^29, 30^. A table describing the metric naming convention can be found in the Supplemental Materials.

**Figure 2:**
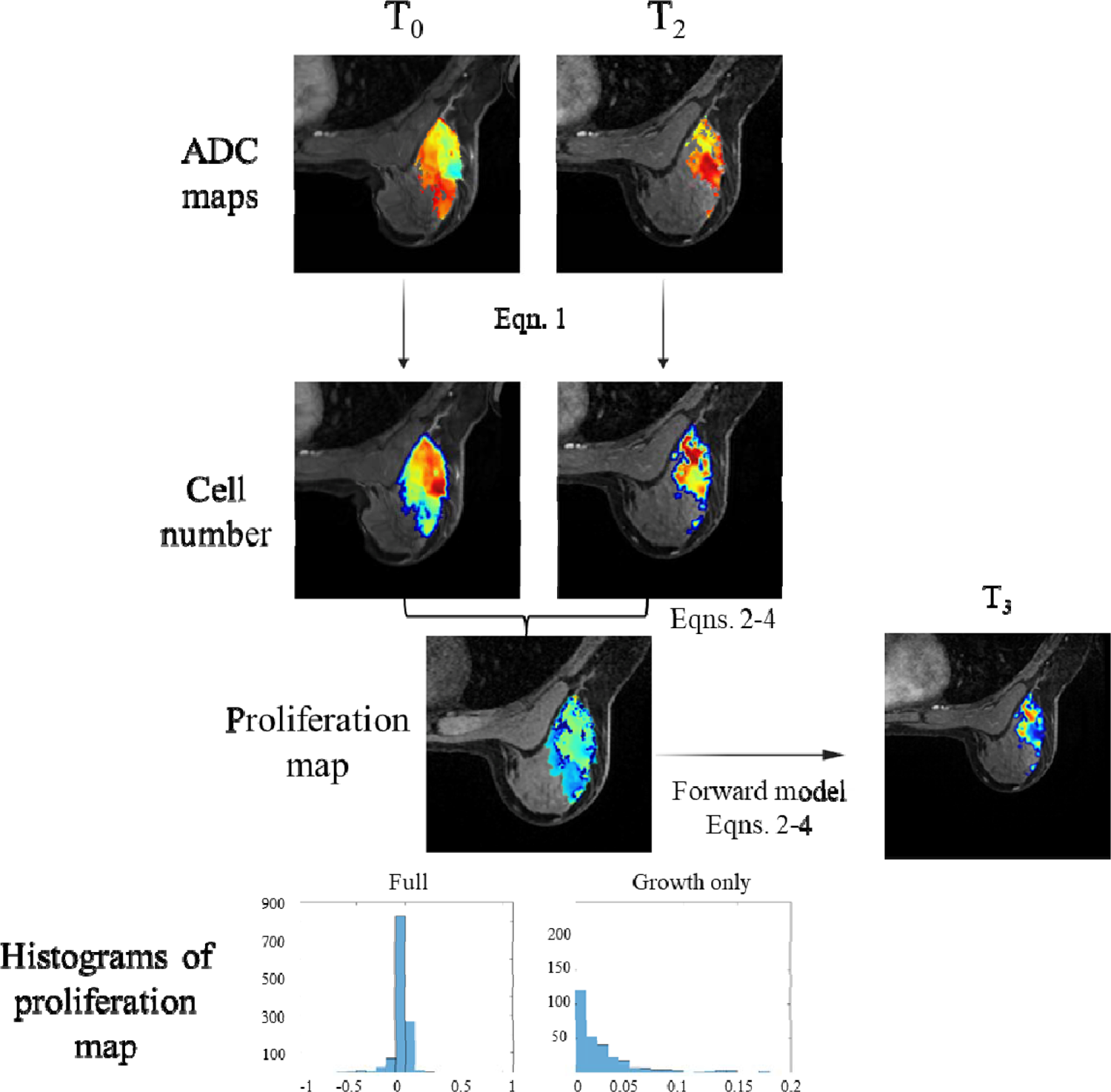
Schematic of the model-based methodology for characterization of NAT response. ADC maps were used to estimate of tumor cellularity at each observed imaging time point. Region-based spatial proliferation maps were estimated between pairs of imaging time points and then estimated using the biophysical model. Full proliferation histograms as well as growth histograms (k > 0) are generated and used to obtain summary metrics. We also generate a T3 model-based prediction using the fit parameters.

### Statistical Analysis

Univariate statistical analysis between non-pCR and pCR patient groups were performed for each evaluation metric. Model-based metrics using *D_0_*, histogram summary metrics of tumor response, and predicted T_3_ area from model fits between T_0_ and T_1_ (T_01_) and T_0_ and T_2_ (T_02_), and conventional radiological metrics of ΔADC and ΔFTV were used to determine univariate metric pCR predictability. Receiver operating characteristic (ROC) curves were constructed and the areas under the ROC curve (AUCs) were calculated. The 95% confidence interval (CI) of the estimated AUC was constructed and sensitivity (or recall), specificity, precision, and accuracy were reported to further summarize the curve and to compare evaluation metrics as classifiers for pCR. With the unbalanced dataset due to observed pCR rates, methods that quantify the ability to identify the number of correct positive predictions (pCR) (precision), an imbalanced classification accuracy metric balancing precision and sensitivity (F_measure_), and the metric’s overall accuracy at predicting response (accuracy) are important to not bias evaluation of performance. Sub-analyses were also performed based on tumor subtype. *P*-values were used to indicate statistical significance.

## Results

Twenty-one (11%) of the 191 patients were excluded due to failure in longitudinal non-rigid image registration. Two subjects (1%) were excluded due to missing or incorrect images. From the 168 evaluable patients, the mean age was 49 years ± 11. The majority of participants had a tumor grade of III (115 [68.4%] of 168) and either HR+/HER2-(76 [45.2%] of 168) or HR-/HER2-(48 [28.6%] of 168) subtypes. After treatment, 31% of the 168 patients had tumors with pCR. Additional demographic data and disease characteristics for the analysis cohort can be found in Table 1.

**Table 1:** Patient demographic data and disease characteristics for analysis cohort.

| **Characteristic** | **Analysis Cohort** |
| --- | --- |
| Mean age ± standard deviation | 49 ± 11 |
| Race |  |
| White | 127 (75.6) |
| Black | 16 (9.5) |
| Asian | 11 (6.6) |
| Other | 1 (0.6) |
| Unknown | 13 (7.7) |
| HR/HER2 subtype |  |
| HR-/HER2- | 48 (28.6) |
| HR+/HER2- | 76 (45.2) |
| HR-/HER2+ | 14 (8.3) |
| HR+/HER2+ | 30 (17.9) |
| Longest Diameter at baseline MRI |  |
| Mean diameter ± standard deviation (cm) | 4.2 ± 2.2 |
| Index lesion type |  |
| Single mass | 65 (38.7) |
| Single NME | 7 (4.2) |
| Multiple masses | 90 (53.5) |
| Multiple NME | 6 (3.6) |
| Tumor grade |  |
| I (Low) | 5 (3) |
| II (Intermediate) | 47 (28) |
| III (High) | 115 (68.4) |
| Missing | 1 (0.6) |

**Table 2:**
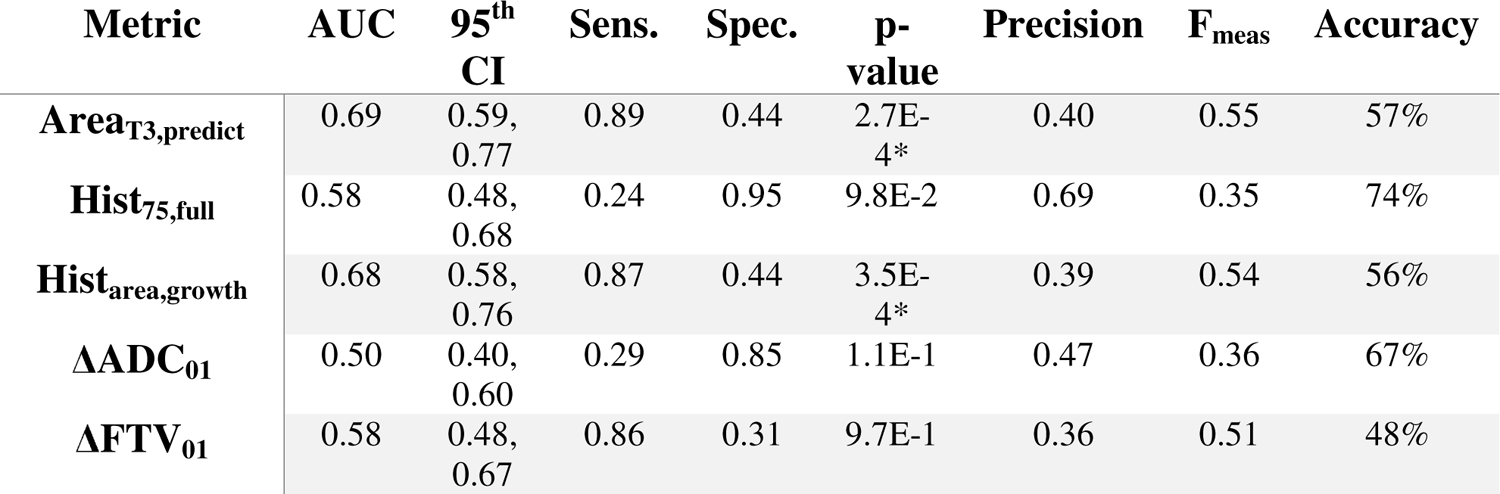
Performance of model-based metrics and conventional radiologic assessment methods for predicting pCR at early treatment (T_01_). * denotes statistical significance (Complete table i located in Supplemental Materials)

| Metric | AUC | 95<sup>th</sup><br>CI | Sens. | Spec. | p-<br>value | Precision | F<sub>meas</sub> | Accuracy |
| --- | --- | --- | --- | --- | --- | --- | --- | --- |
| $\text{Area}_{T3,\text{predict}}$ | 0.69 | 0.59,<br>0.77 | 0.89 | 0.44 | 2.7E-4* | 0.40 | 0.55 | 57% |
| $\text{Hist}_{75,\text{full}}$ | 0.58 | 0.48,<br>0.68 | 0.24 | 0.95 | 9.8E-2 | 0.69 | 0.35 | 74% |
| $\text{Hist}_{\text{area,growth}}$ | 0.68 | 0.58,<br>0.76 | 0.87 | 0.44 | 3.5E-4* | 0.39 | 0.54 | 56% |
| $\Delta\text{ADC}_{01}$ | 0.50 | 0.40,<br>0.60 | 0.29 | 0.85 | 1.1E-1 | 0.47 | 0.36 | 67% |
| $\Delta\text{FTV}_{01}$ | 0.58 | 0.48,<br>0.67 | 0.86 | 0.31 | 9.7E-1 | 0.36 | 0.51 | 48% |

**Table 3:** Performance of model-based metrics and conventional assessment methods for predicting pCR at midtreatment (T_02_). * denotes significance (Complete table is located in Supplemental Materials)

| *Metric* | **AUC** | **95<sup>th</sup>**<br>**CI** | **Sens.** | **Spec.** | **p-**<br>**value** | **Precision** | **F<sub>measure</sub>** | **Accuracy** |
| --- | --- | --- | --- | --- | --- | --- | --- | --- |
| **Area<sub>T3,predict</sub>** | 0.75 | 0.66,<br>0.82 | 0.81 | 0.62 | 1.84E-7* | 0.49 | 0.61 | 68% |
| **Hist<sub>75,full</sub>** | 0.72 | 0.63,<br>0.81 | 0.62 | 0.79 | 3.84E-7* | 0.57 | 0.59 | 74% |
| **Hist<sub>area,growth</sub>** | 0.76 | 0.67,<br>0.83 | 0.83 | 0.59 | 1.27E-7* | 0.48 | 0.61 | 67% |
| **$\Delta\text{ADC}_{02}$** | 0.47 | 0.37,<br>0.56 | 0.27 | 0.81 | 5.06E-1 | 0.38 | 0.31 | 64% |
| **$\Delta\text{FTV}_{02}$** | 0.65 | 0.55,<br>0.74 | 0.52 | 0.80 | 1.59E-3* | 0.54 | 0.53 | 71% |

Mechanistic model-based metrics of response were evaluated using a biophysical model of tumor growth and response to obtain estimates of global diffusion, spatial proliferation rate, and T_3_ predicted area based on changes in observed cellularity both between baseline and early-treatment (T_01_) and baseline and midtreatment (T_02_). This allows for dynamic characterization of patient-specific response throughout the course of NAT. Representative images of model-based metric quantification in study patients exhibiting different tumor responses to treatment are shown in Figures 3 and 4. Figure 3 shows model-based assessment for a representative responder at T_1_, and qualitatively shows minimal response and a model-based prediction of non-pCR. However, Figure 4 shows based on model assessment at T_2_ the same tumor exhibited a robust response and a model-based prediction of pCR. This example is to highlight the importance of serial imaging and dynamic response assessment throughout NAT. To contrast, Figures 5 and 6 show model-based assessment of a representative non-responder at T_1_ and T_2_, respectively. Qualitatively, the proliferation maps show that the representative non-responsive tumor exhibits a heterogeneous response with some initial resistance in the early treatment response assessment.

**Figure 3:**
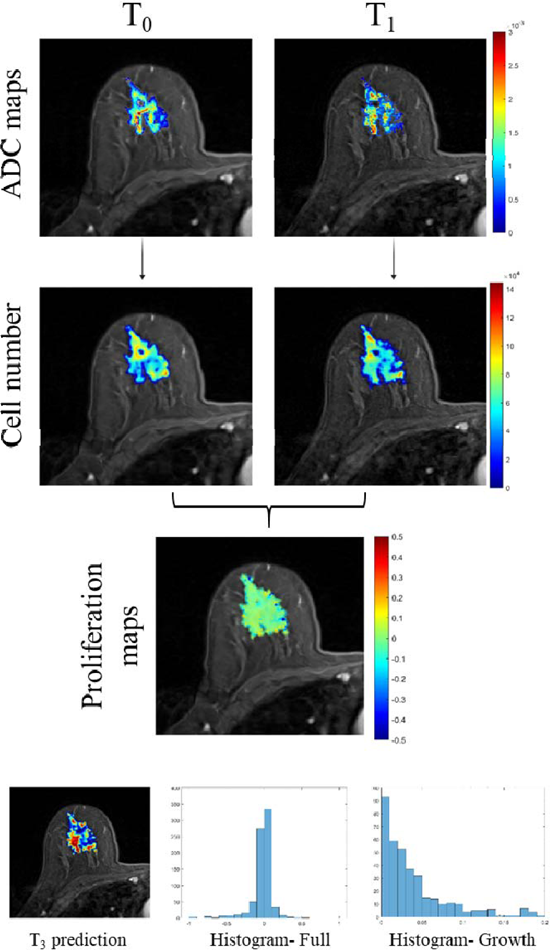
Schematic of early treatment mechanistic modeling methods using T0 and T1 imaging data in a woman who underwent NAT for grade II HR-/HER2+ breast cancer who achieved pCR at surgery. Shown are the patients ADC maps overlaid on their DCE-MR image and the estimated cell number for T_0_ and T_1_. The slices used were selected based on the central slice of the tumor at T_1_. Model-based metrics were as follows: Area_T3,predict_ - 1244 μm2, Hist_75,full_ - 0.028 day-1, Hist_area,growth_ - 38.4. Based on the cutoff values, at T_01_ this tumor would have been predicted non-pCR with the current therapy.

**Figure 4:**
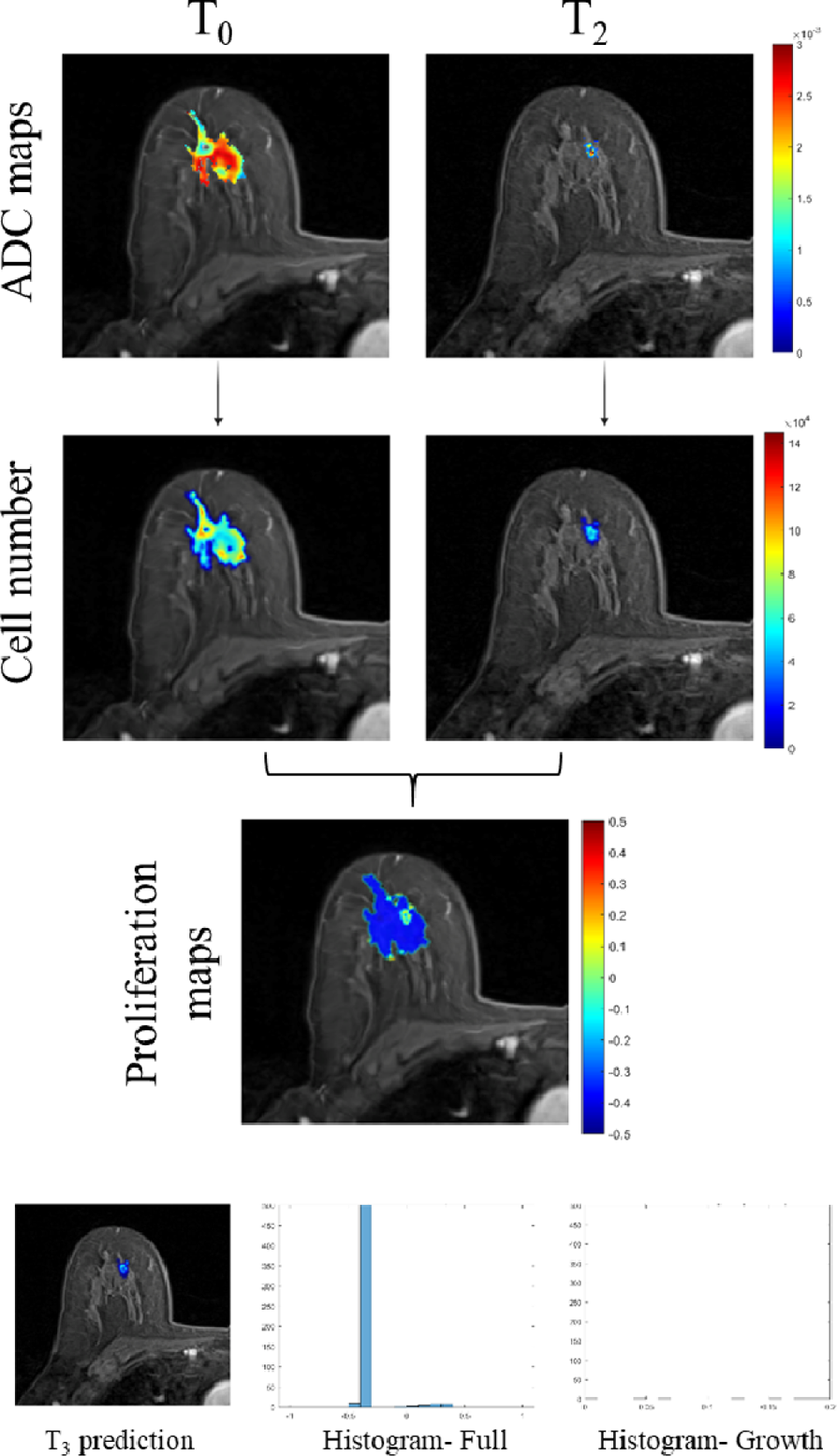
Schematic of midtreatment mechanistic modeling methods using T_0_ and T_2_ in a woman who underwent NAT for grade II HR-/HER2+ breast cancer who had a tumor that achieved pCR. Shown are the ADC maps overlaid on their DCE-MR image and the estimated cell number for T_0_ and T_2_. The slices used were selected based on the central slice of the tumor at T_2_. Model-based metrics were as follows: Area_T3,predict_ - 59 μm^2^, Hist_75,full_ −0.37 day^-1^, Hist_area,growth_ - 2.5. Based on the cutoff values, at T_02_ this tumor would have been predicted pCR.

**Figure 5:**
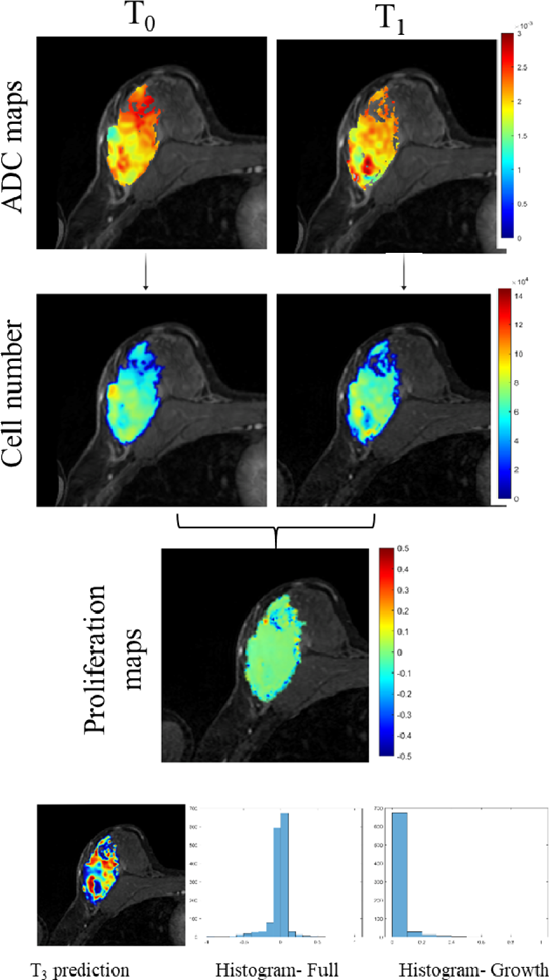
Schematic of early treatment mechanistic modeling methods using T_0_ and T_1_ in a woman who underwent NAT for grade III HR-/HER2-breast cancer who had residual disease at surgery (non-pCR). Shown are the ADC maps overlaid on their DCE-MR image and the estimated cell number for T_0_ and T_1_. The slices used were selected based on the central slice of the tumor at T_1_. Model-based metrics were as follows: Area_T3,predict_ - 2445 μm^2^, Hist_75,full_ - 0.01 day^-1^, Hist_area,growth_ - 71.7. Based on the cutoff values, at T_01_ this tumor would have been predicted as non-pCR.

**Figure 6:**
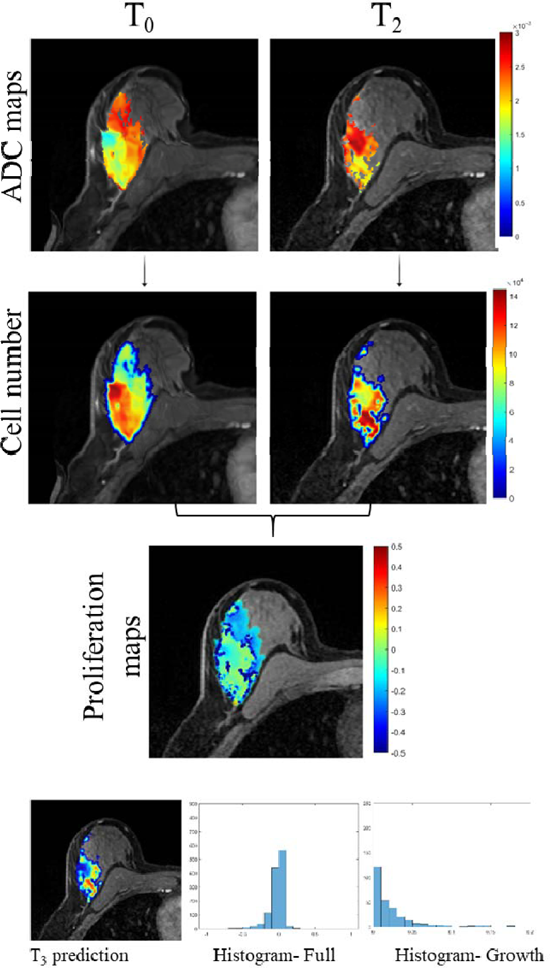
Schematic of midtreatment mechanistic modeling methods using T_0_ and T_2_ in a woman who underwent NAT for grade III HR-/HER2-breast cancer who had residual disease at surgery (non-pCR). Shown are the ADC maps overlaid on their DCE-MR image and the estimated cell number for T_0_ and T_2_. The slices used were selected based on the central slice of the tumor at T_2_. Model-based metrics were as follows: Area_T3,predict_ - 438 μm^2^, Hist_75,full_ −1.3e-3 day^-1^, Hist_area,growth_ - 28.5. Based on the cutoff values, at T_02_ this tumor would have been predicted as non-pCR.

### Early Treatment Response Assessment

ROC results for summary metrics from T_01_ model assessments are detailed in Table 4.2. Model-based summary metrics of Area_T3,predict_ and the Hist_area,growth_ using data T_0_ and T_1_ (T_01_) were found to be statistically significant for differentiating between pCR and non-pCR. Model-based metric of Area_T3,predict_ from T_01_ was found to be statistically significant (*p* < 0.001) with an AUC of 0.69 and with a high sensitivity (0.89); however, the precision for this metric was only 0.4. Hist_75,full_ from T_01_ was not statistically significant (*p* > 0.05) with an AUC of 0.58; however, it was highly specific (0.95), adequate precision (0.69), but had a low sensitivity (0.24). In comparison to conventional radiologic assessment methods of ΔADC and ΔFTV, our model-based metrics show enhanced performance in all statistical performance assessment measures. Figure 7 shows the ROC curves for each reported metric and qualitatively shows that Area_T3,predict_ and the Hist_area,growth_ have enhanced pCR predictability over that of other displayed metrics with clear separation in the ROC curves.

**Figure 7:**
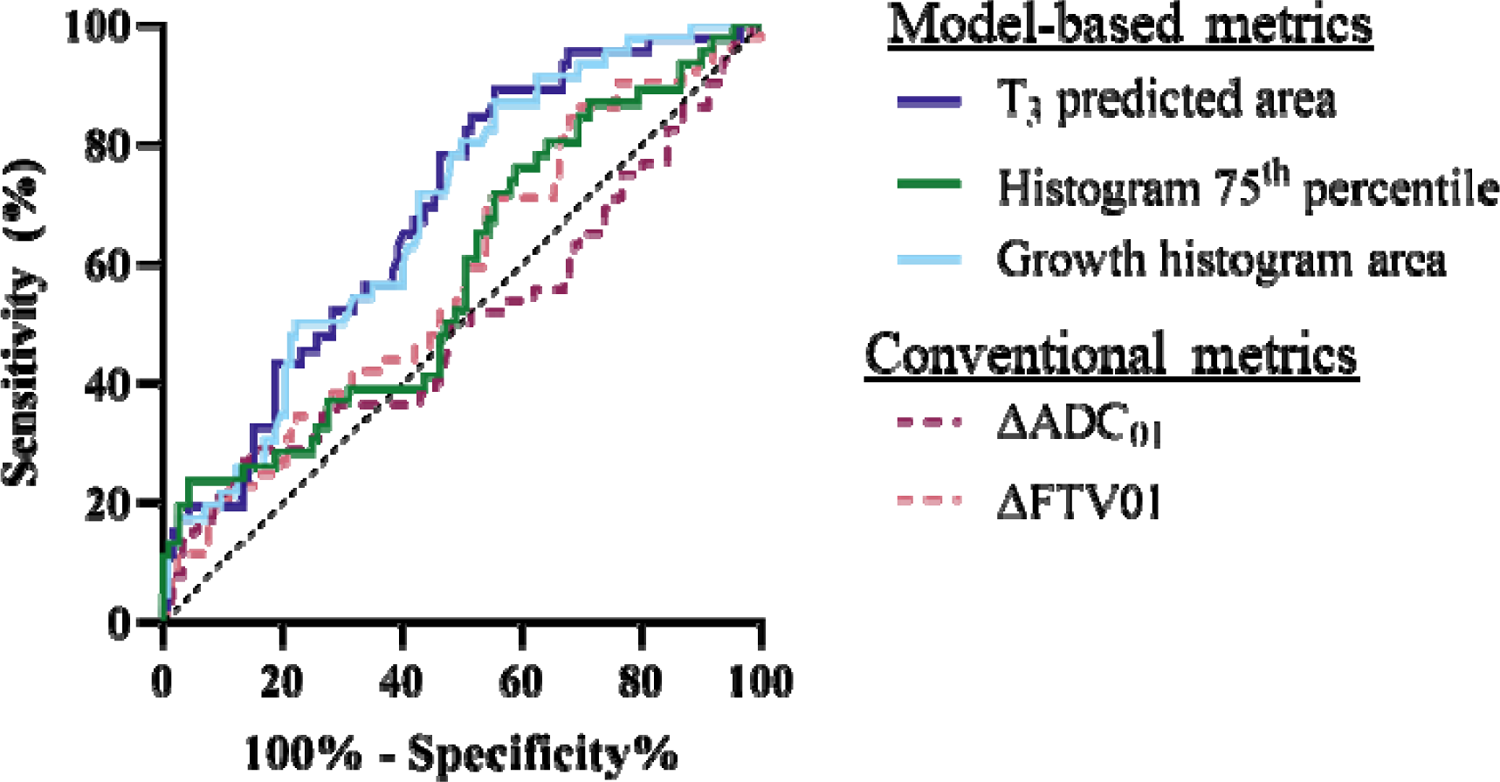
ROC curves for model-based metrics of Area_T3,predict_, Hist_75,full_, and the Hist_area,growth_ to compare to conventional radiologic assessment metrics of change in ADC and change in FTV using early treatment analysis (T_01_).

### Midtreatment Response Assessment

ROC results for summary metrics from baseline and midtreatment time point combinations are detailed in Table 4-3. Model-based summary metrics of Area_T3,predict_, Hist_75,full_, Hist_area,growth_, and ΔFTV using data T_0_ and T_2_ (T_02_) were found to be statistically significant. Model-based metric of Hist_area,growth_ from T_02_ was found to be statistically significant (*p* < 0.001) with an AUC of 0.76, with a high sensitivity (0.83), and moderate specificity (0.59); however, the precision for this metric was only 0.48. Hist_75,full_ was found statistically significant (*p* < 0.001) with an AUC of 0.72, but interestingly was found to be more specific, less sensitive, but more precise when compared to Hist_area,growth_. ΔFTV from T_02_ was statistically significant (*p* < 0.001) with an AUC of 0.65 with moderate sensitivity (0.52), and high specificity (0.80); however, it did have better precision (0.54) when compared to Hist_area,growth_. In comparison, overall our model-based metrics outperform the conventional radiologic assessment methods of ΔADC and ΔFTV for AUC, sensitivity, precision, F_measure_, and accuracy. Figure 8 displays the ROC curves for each reported metric and qualitatively shows that our model-based metrics have enhanced pCR predictability over conventional radiologic metrics.

**Figure 8:**
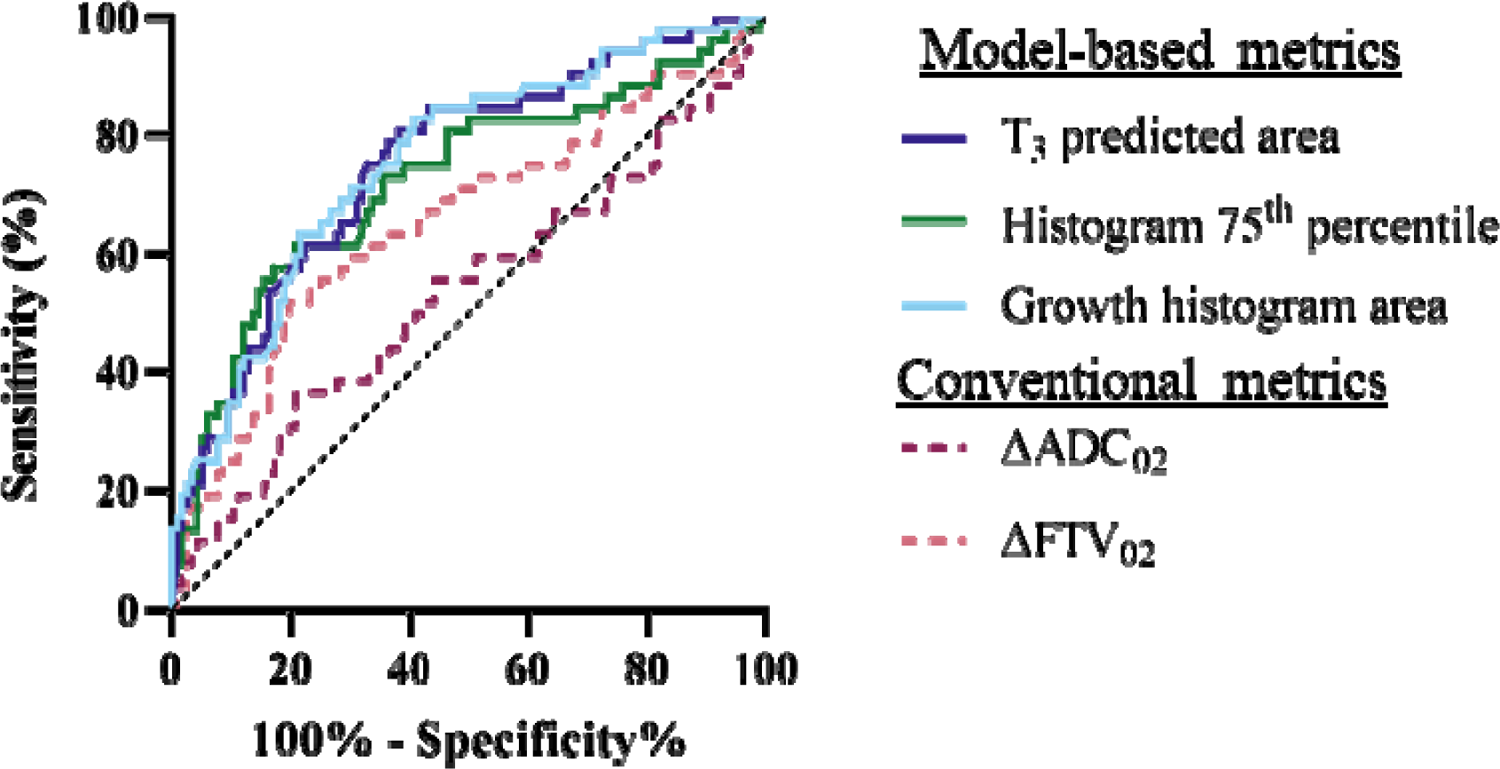
ROC curves for model-based metrics of Area_T3,predict_, Hist_75,full_, and the Hist_area,growth_ to compare to conventional radiologic assessment metrics of change in ADC and change in FTV using midtreatment analysis (T_02_).

### Influence of Lesion Subtype on Predictive Value of Mechanistic Model Measures

Treatment response and pCR rates are known to vary with underlying tumor biology, therefore we explored the predictive value of model-based metrics in post-hoc analysis by breast tumor molecular subtype. Predictive value was found to vary significantly by subtype. ROC curves stratified by molecular subtype for Area_T3,predict_, Hist_75,full_, Hist_area,growth_, ΔADC, and ΔFTV for T_01_ and T_02_ can be found in Figures 9 and 10, respectively. When stratifying by subtype, model-based metrics performed best on the HR-/HER2+ subtype. Figure 9 shows that model-based metrics of Area_T3,predict_ and Hist_area,growth_ achieved AUC of 0.90 at early treatment assessment for HR-/HER2+ disease. Figure 10 shows that model-based metrics of Area_T3,predict_, Hist_75,full,_ and, Hist_area,growth_ classified HR-/HER2+ patients at midtreatment assessment (T_02_) with AUC of 1.0. Summary table of metrics for each ROC curve can be found in Supplemental Materials.

**Figure 9:**
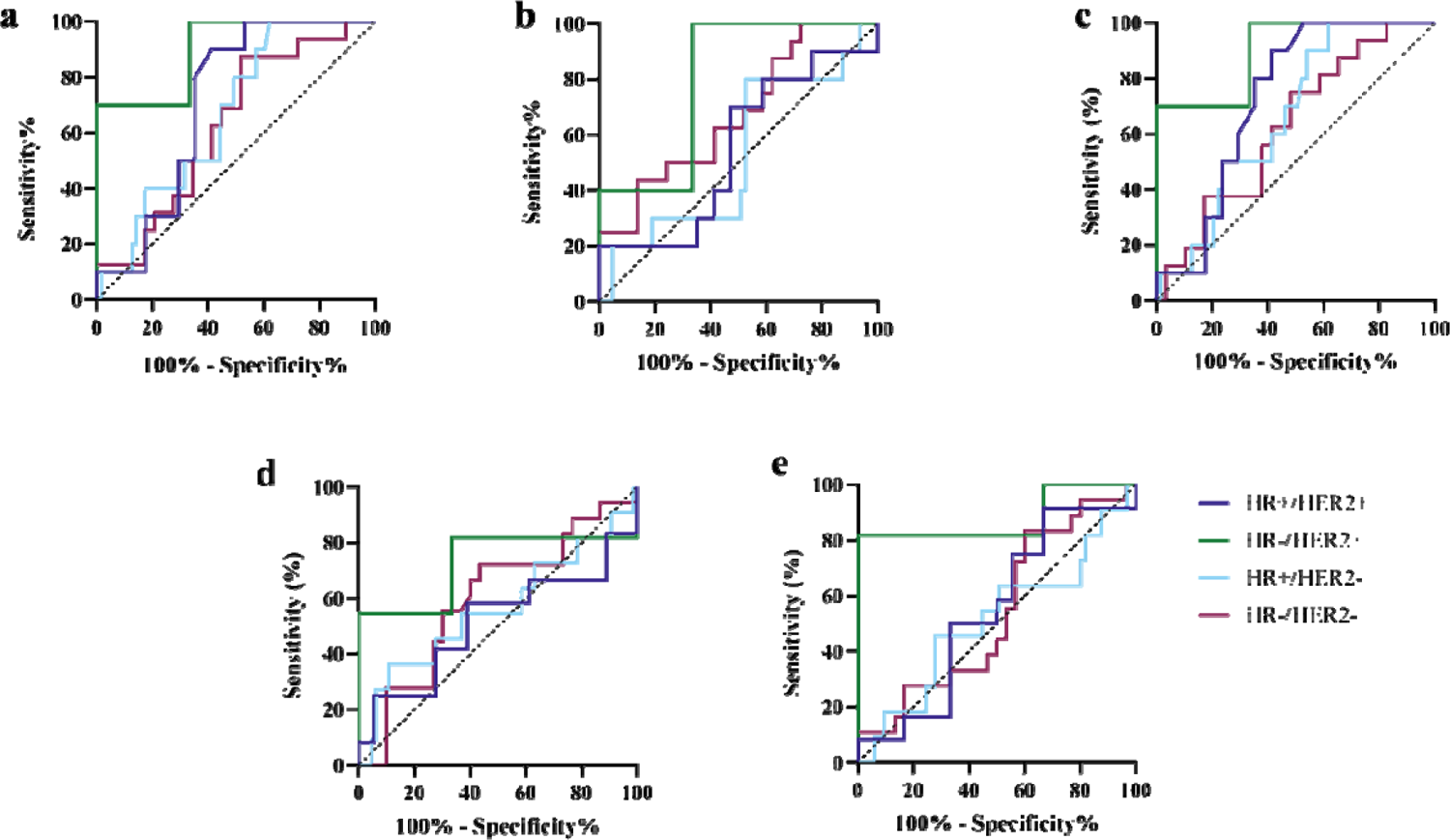
ROC curves for predicting pCR based on early treatment (T_01_) response assessment using a) Area_T3,predict_, b) Hist_75,full_, c), Hist_area,growth_, d) percent change in ADC, and e) percent change in FTV, stratified by tumor subtype.

**Figure 10:**
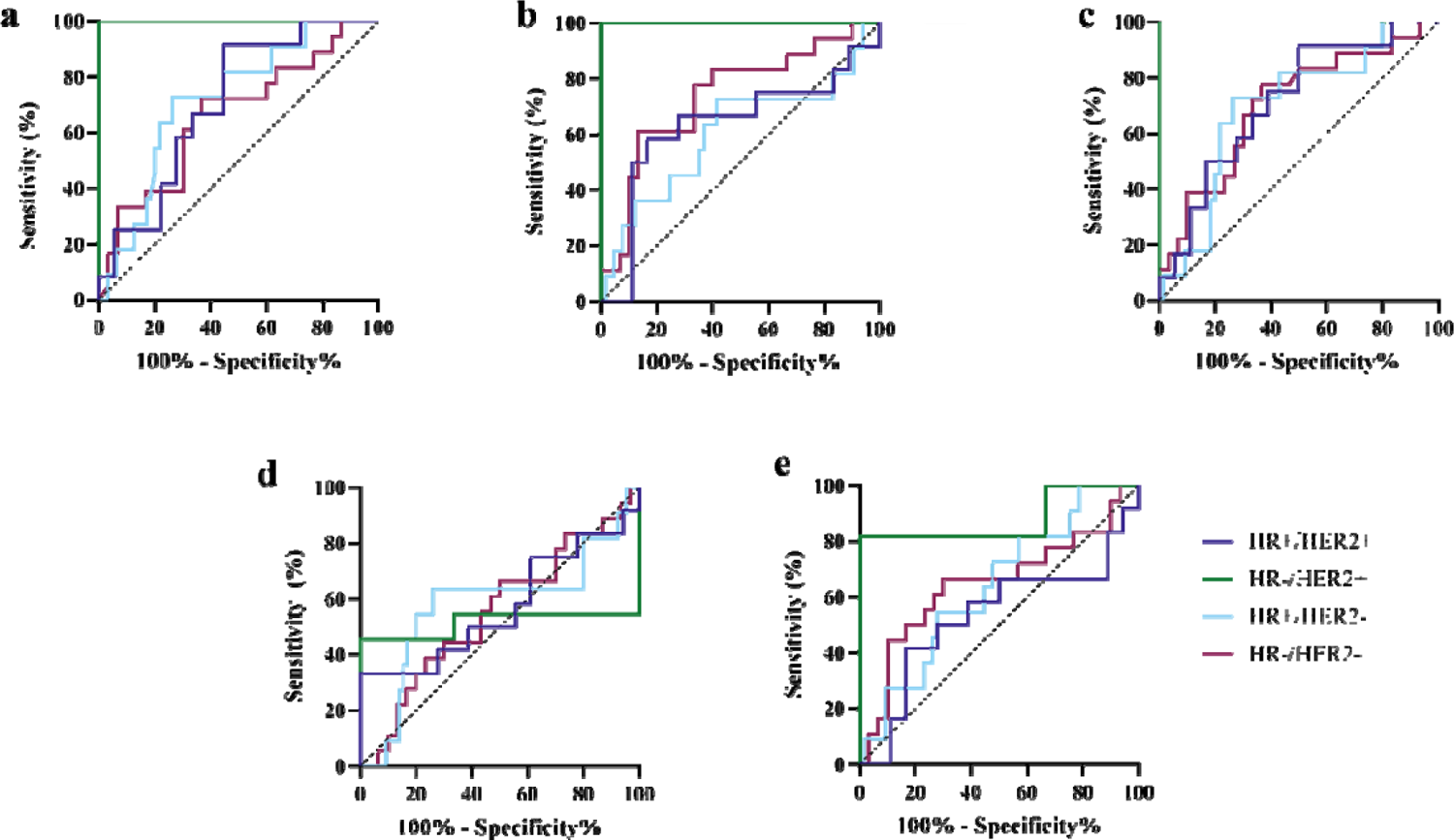
ROC curves for predicting pCR based on midtreatment (T_02_) response assessment using a) Area_T3,predict_, b) Hist_75,full_, c), Hist_area,growth_, d) percent change in ADC, and e) percent change in FTV, stratified by tumor subtype.

## Discussion

Results from this study demonstrate that our mechanistic model-based methods provide a noninvasive and quantitative imaging biomarker to characterize response to NAT for breast cancer and predict pCR. Our study found that model-based metrics at both early and midtreatment time points were predictive of pathologic response with ROC AUC of 0.69 and 0.76, respectively, for the best performing metrics in univariate analysis of 168 breast tumors. Model-estimated proliferation maps from representative responsive and non-responsive tumors highlights how this work could also serve as a qualitative visualization aid to highlight spatial components of response/resistance and heterogeneity that could support in additional clinical decision making for guided biopsies of resistant regions.

As a necessary step towards eventual clinical translation, we sought to examine the multi-site pCR prediction performance of biophysical model-based interpretations of quantitative MRI during the course of breast cancer NAT. The need for multi-site evaluation in this setting is underscored by the often-conflicting nature of imaging-based assessments of therapy response. In much of the literature, imaging biomarker studies lead to conflicting results with a disappointing lack of confirmation when analyses are deployed to different sites with different research teams and different scanner vendors. Significant challenges are thus often expected in generalizing single-center results to the multi-site setting due to inter-site differences in imaging protocol implementations (e.g. varying manufacturers, field strengths, and capabilities) that lead to variable image quality, and differing image reconstruction, scaling, and the generation of quantitative image-based metrics. Our use of a biophysical modeling analysis framework represents a fundamentally different use of imaging data than conventional image-based analysis that strives to overcome these previous challenges, but there is a considerable need for study designs that directly test the hypothesis of biophysical model-based analysis and prediction in realistic clinical trial settings. This study represents an extension to a previous study demonstrating proof-of-concept for the use of a mechanically-coupled reaction-diffusion model for breast cancer NAT treatment response which examined predictions of response outcomes using imaging data from before and after a single cycle of NAT. This previous study was conducted at a single site within the context of a multi-parametric MRI research study with 33 patients enrolled and scanned on dedicated research scanners that achieved AUC for pCR prediction of 0.87^10^. The current work extends this previous study through the expansion to multi-site acquisition on clinical scanners in the context of the I-SPY 2 treatment clinical trial. Our methods offer a significant advantage compared to other conventional radiological response assessment methods as our mechanistic model-based metrics eliminates the site-dependent harmonization of quantitative ADC value standardization to minimize variability between sites, scanners, and patients by using a biophysics-based model to quantify spatiotemporal changes in observed images in an individual patient through time. As expected, we found somewhat lower AUC in this multi-site study as compared to our previous single-site study (0.75 vs. 0.87), we demonstrate the considerable potential for biophysical model-based analysis in realistic clinical trial settings. In future work, we aim to improve predictive capabilities by combining model-based features into multi-parametric signatures of treatment response as well as improving the binary outcome metric of pCR with a continuous outcome metric of pathologic response using more in-depth analysis of residual disease.

Clinical treatment response assessment decision-support tools that can help optimize therapeutic dosing regimens have the potential to support dynamic therapy de-escalation/escalation strategies to avoid unnecessary or ineffective cytotoxic therapies and minimize treatment-related toxicities. However, new methods are needed as current clinical response assessment methods are too coarse to support individualized decisions. In developing new tools for decision-support, it is critical to rigorously examine the association of evaluation metrics with observed outcomes. Historically, ROC AUC, sensitivity, and specificity parameters have been used to evaluate response assessment biomarkers for pCR prediction. While these statistical evaluation measures do well at describing overall classification accuracy, their use in imbalanced outcome datasets leads to biased estimators of performance. pCR rates in this study were 30%, reflecting an imbalanced dataset. A more comprehensive solution that yields greater insight when examining such imbalanced outcomes, is the use of precision and recall statistical measures. Precision is a metric which quantifies the number of positive class (pCR) predictions that actually achieve pCR. High precision is necessary when identifying patients eligible for therapy de-escalation, as identification should be selective with eligibility exclusive to only true responders. In the context of therapy de-escalation, identification of metrics that minimize false positive pCR, while also identifying all true positive pCR patients is desired. Recall, which is calculated similar to sensitivity, quantifies the number of positive pCR predictions made out of all positive pCR patients in the dataset. In a perfect setting, a metric would identify all patients with eventual pCR through early response assessment, however it is important to note that some false negative predictions do not necessarily indicate failure of an early evaluation metric. It is possible that the subsequent additional therapy was required for complete tumor eradication. For these patients, our response assessment methods may be useful in supporting early or midtreatment biopsy assessment of resistant tumor regions, guided from the model-estimated spatiotemporal proliferation maps, to assist clinical decision making as a quantitative assessment of response.

The results of this study are promising but are not without several important limitations. At the time of data retrieval, MRI studies from the conclusion of therapy prior to surgery (T_3_) were unavailable. We show that T_3_ predicted area at early and midtreatment time points is predictive of eventual pathological response, but it is also of interest to compare our T_3_ response predictions to observed T_3_ imaging data to evaluate prediction of radiological response. In general, radiologic complete response has been previously shown as a poor predictor of pathologic complete response, however it has been shown as predictive of recurrence-free survival^31^. As such, it is possible that pCR prediction may not be the only important outcome metric for evaluation of response prediction. In previous work^16^, we describe a framework capable of capturing the dynamic, patient-specific response to NAT with excellent correlation of proliferation histogram summary metrics to residual cancer burden (RCB)^32^ – a continuous metric that is calculated based on components of tumor size, tumor cellularity, and ALN involvement. For a comprehensive analysis in this multi-site dataset it is critical that evaluated outcome metrics complement the observed assessment metrics. This is especially important in the context of isolating differences in complete response and in-breast response, as current methods lack the ability to characterize ALN status. A complete response assessment of pCR for breast cancer patients would include characterization of both the primary breast tumor and ALN as pCR is definition includes the absence of residual tumor in the breast and/or ALN. However, current methods for image-based ALN status are significantly limited^33^. Despite these limitations, our methods allow for mechanistic characterization of dynamic biophysical changes throughout the course of NAT with demonstrated ability to predict response, improving upon current conventional radiological assessment methods.

In conclusion, we demonstrate that an image-driven model-based analysis can characterize biophysical metrics of spatial proliferation to capture dynamic changes in therapeutic response throughout the course of breast cancer NAT using quantitative imaging data. Our results show significant predictive capabilities for pathological response assessed at the conclusion of therapy using model-based metrics evaluated using data acquired during the course of therapy. Our data suggests that imaging-based biophysical modeling approaches may have the potential to support clinical decision making through early indication of response to motivate therapy de-escalation for the minimization of treatment-related toxicities. In addition to providing a quantitative assessment, our methods may also provide additional utility as a quantitative visualization of the dynamic spatiotemporal heterogeneity of treatment response which may be used to aid clinicians in both guiding midtreatment biopsy to areas of active resistance as well as through educating patients on personalized response to motivate future therapeutic decision making. With further investigation, this work has the potential to advance the development of response-adaptive therapeutic regimens whereby regimens are individually designed based on patient-specific mechanistic observations of dynamic response to therapy.

## Data Availability

The datasets used and/or analyzed under this study are publicly available at The Cancer Imaging Archive (https://www.cancerimagingarchive.net/)

## Acknowledgements

National Institutes of Health – National Cancer Institute K25CA204599 and P30CA012197. Wake Forest Baptist Medical Center Comprehensive Cancer Center Signaling and Biotechnology program pilot grant. Williams Family Chair in Breast Oncology. Some figures were created using BioRender.

## References

1. Krzyszczyk P, Acevedo A, Davidoff EJ, Timmins LM, Marrero-Berrios I, Patel M, White C, Lowe C, Sherba JJ, Hartmanshenn C, O’Neill KM, Balter ML, Fritz ZR, Androulakis IP, Schloss RS, Yarmush ML. The growing role of precision and personalized medicine for cancer treatment. Technology (Singap World Sci). 2018;6(3-4):79–100. PubMed PMID: 30713991; PMCID: PMC6352312.

2. Pfohl U, Pflaume A, Regenbrecht M, Finkler S, Graf Adelmann Q, Reinhard C, Regenbrecht CRA, Wedeken L. Precision Oncology Beyond Genomics: The Future Is Here-It Is Just Not Evenly Distributed. Cells. 2021;10(4). PubMed PMID: 33920536; PMCID: PMC8072767.

3. Bozic I, Reiter JG, Allen B, Antal T, Chatterjee K, Shah P, Moon YS, Yaqubie A, Kelly N, Le DT, Lipson EJ, Chapman PB, Diaz LA, Jr., Vogelstein B, Nowak MA. Evolutionary dynamics of cancer in response to targeted combination therapy. Elife. 2013;2:e00747. PubMed PMID: 23805382; PMCID: PMC3691570.

4. Groenendijk FH, Bernards R. Drug resistance to targeted therapies: déjà vu all over again. Mol Oncol. 2014;8(6):1067–83. PubMed PMID: 24910388; PMCID: PMC5528618.

5. Patwardhan GA, Marczyk M, Wali VB, Stern DF, Pusztai L, Hatzis C. Treatment scheduling effects on the evolution of drug resistance in heterogeneous cancer cell populations. NPJ Breast Cancer. 2021;7(1):60. PubMed PMID: 34040000; PMCID: PMC8154902.

6. Kaiser WA, Zeitler E. MR imaging of the breast: fast imaging sequences with and without Gd-DTPA. Preliminary observations. Radiology. 1989;170(3 Pt 1):681-6. PubMed PMID: 2916021.

7. Park SH, Moon WK, Cho N, Song IC, Chang JM, Park IA, Han W, Noh DY. Diffusion-weighted MR imaging: pretreatment prediction of response to neoadjuvant chemotherapy in patients with breast cancer. Radiology. 2010;257(1):56–63. PubMed PMID: 20851939.

8. Newitt DC, Zhang Z, Gibbs JE, Partridge SC, Chenevert TL, Rosen MA, Bolan PJ, Marques HS, Aliu S, Li W, Cimino L, Joe BN, Umphrey H, Ojeda-Fournier H, Dogan B, Oh K, Abe H, Drukteinis J, Esserman LJ, Hylton NM. Test–retest repeatability and reproducibility of ADC measures by breast DWI: Results from the ACRIN 6698 trial. Journal of Magnetic Resonance Imaging. 2019;49(6):1617–28.

9. Alizadeh AA, Aranda V, Bardelli A, Blanpain C, Bock C, Borowski C, Caldas C, Califano A, Doherty M, Elsner M, Esteller M, Fitzgerald R, Korbel JO, Lichter P, Mason CE, Navin N, Pe’er D, Polyak K, Roberts CW, Siu L, Snyder A, Stower H, Swanton C, Verhaak RG, Zenklusen JC, Zuber J, Zucman-Rossi J. Toward understanding and exploiting tumor heterogeneity. Nat Med. 2015;21(8):846–53. PubMed PMID: 26248267; PMCID: PMC4785013.

10. Weis JA, Miga MI, Arlinghaus LR, Li X, Abramson V, Chakravarthy AB, Pendyala P, Yankeelov TE. Predicting the Response of Breast Cancer to Neoadjuvant Therapy Using a Mechanically Coupled Reaction–Diffusion Model. Cancer Res. 2015;75(22):4697–707.

11. Weis JA, Miga MI, Arlinghaus LR, Li X, Chakravarthy AB, Abramson V, Farley J, Yankeelov TE. A mechanically coupled reaction–diffusion model for predicting the response of breast tumors to neoadjuvant chemotherapy. Physics in Medicine and Biology. 2013;58(17):5851–66.

12. Weis JA, Miga MI, Yankeelov TE. Three-dimensional image-based mechanical modeling for predicting the response of breast cancer to neoadjuvant therapy. Comput Meth Appl Mech Eng. 2017;314:494–512.

13. Yankeelov TE, Atuegwu N, Hormuth D, Weis JA, Barnes SL, Miga MI, Rericha EC, Quaranta V. Clinically Relevant Modeling of Tumor Growth and Treatment Response. Sci Transl Med. 2013;5(187):187ps9.

14. Atuegwu NC, Arlinghaus LR, Li X, Chakravarthy AB, Abramson VG, Sanders ME, Yankeelov TE. Parameterizing the Logistic Model of Tumor Growth by DW-MRI and DCE-MRI Data to Predict Treatment Response and Changes in Breast Cancer Cellularity during Neoadjuvant Chemotherapy. Transl Oncol. 2013;6(3):256–64.

15. Jarrett AM, Hormuth DA, Wu C, Kazerouni AS, Ekrut DA, Virostko J, Sorace AG, DiCarlo JC, Kowalski J, Patt D, Goodgame B, Avery S, Yankeelov TE. Evaluating patient-specific neoadjuvant regimens for breast cancer via a mathematical model constrained by quantitative magnetic resonance imaging data. Neoplasia. 2020;22(12):820–30.

16. Bowers HJ, Douglas E, Ansley K, Thomas A, Weis JA. Dynamic characterization of breast cancer response to neoadjuvant therapy using biophysical metrics of spatial proliferation. Sci Rep. 2022;12(1):11718. PubMed PMID: 35810187; PMCID: PMC9271064.

17. Jenkinson M, Bannister P, Brady M, Smith S. Improved optimization for the robust and accurate linear registration and motion correction of brain images. NeuroImage. 2002;17(2):825–41.

18. Jenkinson M, Smith S. A global optimisation method for robust affine registration of brain images. Medical Image Analysis. 2001;5(2):143–56.

19. Greve DN, Fischl B. Accurate and robust brain image alignment using boundary-based registration. NeuroImage. 2009;48(1):63–72.

20. Ou Y, Sotiras A, Paragios N, Davatzikos C. DRAMMS: Deformable registration via attribute matching and mutual-saliency weighting. Medical Image Analysis. 2011;15(4):622–39.

21. Li X, Arlinghaus LR, Ayers GD, Chakravarthy AB, Abramson RG, Abramson VG, Atuegwu N, Farley J, Mayer IA, Kelley MC, Meszoely IM, Means-Powell J, Grau AM, Sanders M, Bhave SR, Yankeelov TE. DCE-MRI analysis methods for predicting the response of breast cancer to neoadjuvant chemotherapy: pilot study findings. Magn Reson Med. 2014;71(4):1592–602.

22. Atuegwu NC, Colvin DC, Loveless ME, Xu L, Gore JC, Yankeelov TE. Incorporation of diffusion-weighted magnetic resonance imaging data into a simple mathematical model of tumor growth. Physics in Medicine and Biology. 2012;57(1):225–40.

23. Anderson A, Xie J, Pizzonia J, Bronen R, Spencer DD, Gore JC. Effect of cell volume fraction changes on apparent diffusion in human cells. Magnetic Resonance Imaging. 2000;18:689–95.

24. Martin I, Dozin B, Quarto R, Cancedda R, Beltrame F. Computer-based technique for cell aggregation analysis and cell aggregation in in vitro chondrogenesis. Cytometry. 1997;28(2):141–6. PubMed PMID: 9181304.

25. Garg I, Miga MI, editors. Preliminary investigation of the inhibitory effects of mechanical stress in tumor growth. Medical Imaging 2008: Visualization, Image-Guided Procedures, and Modeling; 2008 2008/03/17/: International Society for Optics and Photonics.

26. Liu DC, Nocedal J. On the limited memory BFGS method for large scale optimization. Mathematical Programming. 1989;45(1-3):503–28.

27. Lions JL, Magenes E. Non-Homogeneous Boundary Value Problems and Applications: Vol. 1: Springer Science & Business Media; 2012 2012/12/06/. 375 p.

28. Partridge SC, Zhang Z, Newitt DC, Gibbs JE, Chenevert TL, Rosen MA, Bolan PJ, Marques HS, Romanoff J, Cimino L, Joe BN, Umphrey HR, Ojeda-Fournier H, Dogan B, Oh K, Abe H, Drukteinis JS, Esserman LJ, Hylton NM. Diffusion-weighted MRI Findings Predict Pathologic Response in Neoadjuvant Treatment of Breast Cancer: The ACRIN 6698 Multicenter Trial. Radiology. 2018;289(3):618–27.

29. Hylton NM. Vascularity assessment of breast lesions with gadolinium-enhanced MR imaging. Magn Reson Imaging Clin N Am. 1999;7(2):411–20, x. PubMed PMID: 10382170.

30. Hylton NM, Gatsonis CA, Rosen MA, Lehman CD, Newitt DC, Partridge SC, Bernreuter WK, Pisano ED, Morris EA, Weatherall PT, Polin SM, Newstead GM, Marques HS, Esserman LJ, Schnall MD. Neoadjuvant Chemotherapy for Breast Cancer: Functional Tumor Volume by MR Imaging Predicts Recurrence-free Survival—Results from the ACRIN 6657/CALGB 150007 I-SPY 1 TRIAL. Radiology. 2016;279(1):44–55.

31. Gampenrieder SP, Peer A, Weismann C, Meissnitzer M, Rinnerthaler G, Webhofer J, Westphal T, Riedmann M, Meissnitzer T, Egger H, Klaassen Federspiel F, Reitsamer R, Hauser-Kronberger C, Stering K, Hergan K, Mlineritsch B, Greil R. Radiologic complete response (rCR) in contrast-enhanced magnetic resonance imaging (CE-MRI) after neoadjuvant chemotherapy for early breast cancer predicts recurrence-free survival but not pathologic complete response (pCR). Breast Cancer Research. 2019;21(1):19.

32. Symmans WF, Wei C, Gould R, Yu X, Zhang Y, Liu M, Walls A, Bousamra A, Ramineni M, Sinn B, Hunt K, Buchholz TA, Valero V, Buzdar AU, Yang W, Brewster AM, Moulder S, Pusztai L, Hatzis C, Hortobagyi GN. Long-Term Prognostic Risk After Neoadjuvant Chemotherapy Associated With Residual Cancer Burden and Breast Cancer Subtype. J Clin Oncol. 2017;35(10):1049–60. PubMed PMID: 28135148; PMCID: PMC5455352.

33. Rahbar H, Conlin JL, Parsian S, DeMartini WB, Peacock S, Lehman CD, Partridge SC. Suspicious axillary lymph nodes identified on clinical breast MRI in patients newly diagnosed with breast cancer: can quantitative features improve discrimination of malignant from benign? Acad Radiol. 2015;22(4):430–8. PubMed PMID: 25491740; PMCID: PMC4355079.

34. Clark K, Vendt B, Smith K, Freymann J, Kirby J, Koppel P, Moore S, Phillips S, Maffitt D, Pringle M, Tarbox L, Prior F. The Cancer Imaging Archive (TCIA): maintaining and operating a public information repository. J Digit Imaging. 2013;26(6):1045–57. PubMed PMID: 23884657; PMCID: PMC3824915.

