## Supplemental Material for "Characterizing breast cancer response to neoadjuvant therapy based on biophysical modeling and multiparametric magnetic resonance imaging data"

##### **Supplemental Materials**

**Table S1:** Full list of univariate metrics analyzed, their description, and the performance of model-based metrics and conventional radiological assessment methods for predicting pCR at early treatment (T_01_).

| **Metric** | **Metric description** | **AUC** | **95% CI** | **P** | **Sens** | **Spec** | **Precision** | **F_m_** | **Accuracy** |
| --- | --- | --- | --- | --- | --- | --- | --- | --- | --- |
| **D_0_** | Cell diffusion in absence of stress | 0.54 | 0.44, 0.64 | 3.9E-01 | 0.65 | 0.46 | 0.33 | 0.44 | 0.52 |
| **Hist_area,Full_** | Histogram area - Full | 0.66 | 0.57, 0.75 | 1.4E-03 | 0.91 | 0.35 | 0.37 | 0.53 | 0.52 |
| **Hist_area,growth_** | Histogram area - Growth | 0.68 | 0.59, 0.76 | 3.5E-04 | 0.87 | 0.44 | 0.39 | 0.54 | 0.57 |
| **Hist_10,full_** | Histogram 10th percentile - Full | 0.54 | 0.44, 0.65 | 4.1E-01 | 0.26 | 0.86 | 0.43 | 0.32 | 0.68 |
| **Hist_10,growth_** | Histogram 10th percentile - Growth | 0.55 | 0.45, 0.64 | 3.3E-01 | 0.69 | 0.48 | 0.36 | 0.47 | 0.54 |
| **Hist_25,full_** | Histogram 25th percentile - Full | 0.56 | 0.45, 0.66 | 2.3E-01 | 0.54 | 0.68 | 0.41 | 0.47 | 0.64 |
| **Hist_25,growth_** | Histogram 25th percentile - Growth | 0.54 | 0.45, 0.64 | 4.1E-01 | 0.67 | 0.52 | 0.37 | 0.48 | 0.57 |
| **Hist_75,full_** | Histogram 75th percentile - Full | 0.58 | 0.48, 0.68 | 9.8E-02 | 0.24 | 0.95 | 0.69 | 0.35 | 0.75 |
| **Hist_75,growth_** | Histogram 75th percentile - Growth | 0.50 | 0.41, 0.60 | 9.8E-01 | 0.89 | 0.23 | 0.33 | 0.48 | 0.43 |
| **Hist_90,full_** | Histogram 90th percentile - Full | 0.55 | 0.46, 0.65 | 3.2E-01 | 0.87 | 0.32 | 0.34 | 0.49 | 0.48 |
| **Hist_90,growth_** | Histogram 90th percentile - Growth | 0.51 | 0.42, 0.61 | 9.1E-01 | 0.69 | 0.42 | 0.33 | 0.45 | 0.50 |
| **Hist_IQR,full_** | Histogram Interquartile Range - Full | 0.50 | 0.42, 0.60 | 9.3E-01 | 0.98 | 0.14 | 0.32 | 0.48 | 0.39 |
| **Hist_IQR,growth_** | Histogram Interquartile Range - Growth | 0.52 | 0.42, 0.61 | 6.9E-01 | 0.91 | 0.23 | 0.33 | 0.48 | 0.43 |
| **Hist_kurt,full_** | Histogram Kurtosis - Full | 0.54 | 0.44, 0.64 | 4.3E-01 | 0.87 | 0.24 | 0.32 | 0.47 | 0.43 |
| **Hist_kurt,growth_** | Histogram Kurtosis - Growth | 0.53 | 0.42, 0.63 | 5.7E-01 | 0.33 | 0.78 | 0.40 | 0.37 | 0.66 |
| **Hist_mean,full_** | Histogram mean - Full | 0.60 | 0.49, 0.70 | 5.7E-02 | 0.50 | 0.80 | 0.51 | 0.51 | 0.71 |
| **Hist_mean,growth_** | Histogram mean - Growth | 0.50 | 0.41, 0.60 | 9.9E-01 | 0.78 | 0.34 | 0.33 | 0.46 | 0.47 |
| **Hist_med,full_** | Histogram median - Full | 0.61 | 0.50, 0.71 | 3.4E-02 | 0.52 | 0.76 | 0.47 | 0.49 | 0.69 |
| **Hist_med,growth_** | Histogram median - Growth | 0.49 | 0.40, 0.59 | 8.7E-01 | 0.87 | 0.27 | 0.33 | 0.48 | 0.45 |
| **Hist_skew,full_** | Histogram skewness - Full | 0.52 | 0.41, 0.62 | 7.7E-01 | 0.54 | 0.58 | 0.35 | 0.42 | 0.57 |
| **Hist_skew,growth_** | Histogram skewness - Growth | 0.52 | 0.42, 0.62 | 6.7E-01 | 0.33 | 0.77 | 0.39 | 0.37 | 0.65 |
| **Hist_STD,full_** | Histogram Standard Deviation - Full | 0.52 | 0.43, 0.63 | 6.6E-01 | 0.74 | 0.36 | 0.32 | 0.45 | 0.47 |
| **Hist_STD,growth_** | Histogram Standard Deviation - Growth | 0.54 | 0.44, 0.63 | 3.9E-01 | 0.96 | 0.23 | 0.34 | 0.50 | 0.44 |
| **Area_T2,Predict_** | T_2_ predicted area | 0.69 | 0.59, 0.77 | 2.4E-04 | 0.80 | 0.50 | 0.40 | 0.53 | 0.59 |
| **Area_T3,Predict_** | T_3_ predicted area | 0.69 | 0.59, 0.77 | 2.7E-04 | 0.89 | 0.44 | 0.40 | 0.55 | 0.57 |
| **ΔFTV** | Change in Functional tumor volume | 0.50 | 0.40, 0.60 | 9.7E-01 | 0.29 | 0.85 | 0.47 | 0.36 | 0.68 |
| **ΔADC** | Change in apparent diffusion coefficient | 0.58 | 0.47, 0.66 | 1.1E-01 | 0.87 | 0.31 | 0.36 | 0.51 | 0.48 |

**Table S-2:** Full list of univariate metrics analyzed, their description, and the performance of model-based metrics and conventional radiological assessment methods for predicting pCR at midtreatment (T_02_).

| Metric | Metric description | AUC | 95% CI | P-value | Sens | Spec | Precision | F_meas_ | Accuracy |
| --- | --- | --- | --- | --- | --- | --- | --- | --- | --- |
| D_0_ | Cell diffusion in absence of stress | 0.63 | 0.53, 0.72 | 7.80E-03 | 0.56 | 0.75 | 0.50 | 0.53 | 0.69 |
| Hist_area,full_ | Histogram area - Full | 0.69 | 0.60, 0.77 | 1.20E-04 | 0.75 | 0.56 | 0.43 | 0.55 | 0.62 |
| Hist_area,growth_ | Histogram area - Growth | 0.76 | 0.67, 0.83 | 1.27E-07 | 0.83 | 0.59 | 0.48 | 0.61 | 0.67 |
| Hist_10,full_ | Histogram 10th percentile - Full | 0.68 | 0.59, 0.77 | 1.53E-04 | 0.77 | 0.56 | 0.44 | 0.56 | 0.63 |
| Hist_10,growth_ | Histogram 10th percentile - Growth | 0.65 | 0.56, 0.75 | 1.39E-03 | 0.85 | 0.44 | 0.40 | 0.55 | 0.57 |
| Hist_25,full_ | Histogram 25th percentile - Full | 0.69 | 0.59, 0.78 | 1.13E-04 | 0.65 | 0.74 | 0.53 | 0.59 | 0.71 |
| Hist_25,growth_ | Histogram 25th percentile - Growth | 0.64 | 0.54, 0.74 | 3.26E-03 | 0.87 | 0.38 | 0.38 | 0.53 | 0.53 |
| Hist_75,full_ | Histogram 75th percentile - Full | 0.72 | 0.63, 0.81 | 3.84E-06 | 0.62 | 0.79 | 0.57 | 0.59 | 0.74 |
| Hist_75,growth_ | Histogram 75th percentile - Growth | 0.62 | 0.52, 0.71 | 1.32E-02 | 0.67 | 0.55 | 0.40 | 0.50 | 0.59 |
| Hist_90,full_ | Histogram 90th percentile - Full | 0.37 | 0.28, 0.48 | 8.20E-03 | 0.04 | 0.99 | 0.67 | 0.07 | 0.70 |
| Hist_90,growth_ | Histogram 90th percentile - Growth | 0.60 | 0.50, 0.69 | 4.00E-02 | 0.29 | 0.95 | 0.71 | 0.41 | 0.74 |
| Hist_iqr,full_ | Histogram Interquartile Range - Full | 0.60 | 0.50, 0.70 | 3.44E-02 | 0.50 | 0.73 | 0.46 | 0.48 | 0.66 |
| Hist_iqr,growth_ | Histogram Interquartile Range - Growth | 0.57 | 0.47, 0.66 | 1.49E-01 | 0.25 | 0.92 | 0.59 | 0.35 | 0.71 |
| Hist_kurt,full_ | Histogram Kurtosis - Full | 0.49 | 0.40, 0.59 | 7.96E-01 | 0.35 | 0.70 | 0.34 | 0.34 | 0.59 |
| Hist_kurt,growth_ | Histogram Kurtosis - Growth | 0.71 | 0.62, 0.79 | 1.05E-05 | 0.54 | 0.84 | 0.61 | 0.57 | 0.75 |
| Hist_mean,full_ | Histogram mean - Full | 0.69 | 0.60, 0.78 | 5.84E-05 | 0.73 | 0.63 | 0.47 | 0.57 | 0.66 |
| Hist_mean,growth_ | Histogram mean - Growth | 0.62 | 0.51 | 1.10E-02 | 0.71 | 0.54 | 0.41 | 0.52 | 0.60 |
| Hist_med,full_ | Histogram median - Full | 0.68 | 0.58 | 1.96E-04 | 0.62 | 0.76 | 0.53 | 0.57 | 0.71 |
| Hist_med,growth_ | Histogram median - Growth | 0.65 | 0.55 | 2.10E-03 | 0.46 | 0.83 | 0.55 | 0.50 | 0.71 |
| Hist_skew,full_ | Histogram skewness - Full | 0.63 | 0.53 | 8.63E-03 | 0.38 | 0.85 | 0.54 | 0.45 | 0.71 |
| Hist_skew,growth_ | Histogram skewness - Growth | 0.72 | 0.63 | 4.53E-06 | 0.62 | 0.78 | 0.55 | 0.58 | 0.73 |
| Hist_std,full_ | Histogram Standard Deviation - Full | 0.55 | 0.45 | 3.48E-01 | 0.50 | 0.66 | 0.40 | 0.44 | 0.61 |
| Hist_std,growth_ | Histogram Standard Deviation - Growth | 0.54 | 0.42 | 4.29E-01 | 0.29 | 0.89 | 0.54 | 0.38 | 0.70 |
| Area_t2,predict_ | T_3_ predicted area | 0.75 | 0.66 | 1.84E-07 | 0.81 | 0.62 | 0.49 | 0.61 | 0.68 |
| Area_t3,predict_ | Change in Functional tumor volume | 0.65 | 0.55 | 1.59E-03 | 0.52 | 0.80 | 0.54 | 0.53 | 0.71 |
| Δftv | Change in apparent diffusion coefficient | 0.47 | 0.37 | 5.06E-01 | 0.27 | 0.81 | 0.38 | 0.31 | 0.64 |

**Table S3:** Summary table of metrics based on molecular subtype for T_01_ model fits and other conventional radiological metrics.

| **Metric** | **Subtype** | **AUC** | **CI** | **Sens** | **Spec** | ***p* value** | **Prec.** | **F_measure_** | **Acc.** |
| --- | --- | --- | --- | --- | --- | --- | --- | --- | --- |
| T3 pred. area | HR+/HER2+ | 0.71 | 0.48, 0.88 | 0.90 | 0.59 | 7.9E-2 | 0.56 | 0.69 | 70% |
|  | HR-/HER2+ | 0.90 | 0.45, 1.0 | 0.70 | 1.0 | 4.9E-2 | 1.0 | 0.82 | 77% |
|  | HR+/HER2- | 0.67 | 0.32, 0.73 | 1.0 | 0.38 | 9.5E-2 | 0.20 | 0.34 | 47% |
|  | HR-/HER2- | 0.63 | 0.46, 0.79 | 0.88 | 0.48 | 1.7E-1 | 0.48 | 0.62 | 62% |
| Hist. 75^th^ | HR+/HER2+ | 0.55 | 0.31, 0.77 | 0.70 | 0.53 | 7.1E-1 | 0.47 | 0.56 | 59% |
|  | HR-/HER2+ | 0.80 | 0.13, 1.0 | 1.0 | 0.67 | 1.6E-1 | 0.91 | 0.95 | 92% |
|  | HR+/HER2- | 0.53 | 0.32, 0.73 | 0.80 | 0.48 | 7.7E-1 | 0.20 | 0.31 | 52% |
|  | HR-/HER2- | 0.67 | 0.48, .082 | 0.44 | 0.86 | 5.9E-2 | 0.64 | 0.52 | 71% |
| Hist. growth AUC | HR+/HER2+ | 0.73 | 0.47, 0.88 | 0.90 | 0.59 | 5.3E-2 | 0.56 | 0.59 | 70% |
|  | HR-/HER2+ | 0.90 | 0.46, 1.0 | 0.70 | 1.0 | 4.9E-2 | 1.0 | 0.82 | 77% |
|  | HR+/HER2- | 0.66 | 0.50, 0.81 | 1.0 | 0.38 | 9.8E-2 | 0.20 | 0.34 | 47% |
|  | HR-/HER2- | 0.63 | 0.44, 0.78 | 0.75 | 0.52 | 1.7E-1 | 0.46 | 0.57 | 60% |

| **Metric** | **Subtype** | **AUC** | **CI** | **Sens** | **Spec** | ***p* value** | **Prec.** | **F_measure_** | **Acc.** |
| --- | --- | --- | --- | --- | --- | --- | --- | --- | --- |
| ΔADC_01_ | HR+/HER2+ | 0.51 | 0.28, 0.74 | 0.58 | 0.61 | 9.2E-1 | 0.50 | 0.54 | 60% |
|  | HR-/HER2+ | 0.73 | 0.37, 0.94 | 0.55 | 1.0 | 2.8E-1 | 1.0 | 0.71 | 64% |
|  | HR+/HER2- | 0.44 | 0.24, 0.68 | 0.18 | 0.90 | 5.3E-1 | 0.20 | 0.19 | 78% |
|  | HR-/HER2- | 0.40 | 0.23, 0.57 | 1.0 | 0.10 | 2.6E-1 | 0.40 | 0.57 | 44% |
| ΔFTV_01_ | HR+/HER2+ | 0.45 | 0.24, 0.66 | 0.83 | 0.33 | 6.9E-1 | 0.45 | 0.59 | 53% |
|  | HR-/HER2+ | 0.88 | 0.55, 1.0 | 0.82 | 1.0 | 6.0E-2 | 1.0 | 0.9 | 86% |
|  | HR+/HER2- | 0.49 | 0.29, 0.70 | 0.36 | 0.80 | 9.1E-1 | 0.24 | 0.29 | 74% |
|  | HR-/HER2- | 0.54 | 0.38, 0.71 | 0.83 | 0.40 | 6.5E-1 | 0.45 | 0.59 | 56% |

**Table S4:** Summary table of metrics based on molecular subtype for T_02_ model fits and other conventional radiological metrics.

| **Metric** | **Subtype** | **AUC** | **CI** | **Sens** | **Spec** | ***p* value** | **Prec** | **F_meas_** | **Acc.** |
| --- | --- | --- | --- | --- | --- | --- | --- | --- | --- |
| T3 pred. area | HR+/HER2+ | 0.71 | 0.49, 0.88 | 0.92 | 0.56 | 6.0E-02 | 0.58 | 0.71 | 70% |
|  | HR-/HER2+ | 1.0 | 1.0, 1.0 | 1.0 | 1.0 | 5.5E-3* | 1.0 | 1.0 | 100% |
|  | HR+/HER2- | 0.72 | 0.53, 0.85 | 0.73 | 0.74 | 1.9E-2* | 0.32 | 0.44 | 74% |
|  | HR-/HER2- | 0.66 | 0.49, 0.81 | 0.72 | 0.63 | 6.0E-2 | 0.54 | 0.62 | 67% |
| Hist. 75^th^ | HR+/HER2+ | 0.63 | 0.39, 0.85 | 0.58 | 0.83 | 2.3E-01 | 0.70 | 0.64 | 73% |
|  | HR-/HER2+ | 1.0 | 1.0, 1.0 | 1.0 | 1.0 | 5.5E-3* | 1.0 | 1.0 | 100% |
|  | HR+/HER2- | 0.61 | 0.34, 0.79 | 0.73 | 0.58 | 2.6E-1 | 0.23 | 0.35 | 61% |
|  | HR-/HER2- | 0.74 | 0.54, 0.87 | 0.61 | 0.87 | 6.2E-3* | 0.73 | 0.67 | 77% |
| Hist. growth AUC | HR+/HER2+ | 0.71 | 0.48, 0.87 | 0.92 | 0.50 | 5.4E-02 | 0.55 | 0.69 | 67% |
|  | HR-/HER2+ | 1.0 | 1.0, 1.0 | 1.0 | 1.0 | 5.5E-3* | 1.0 | 1.0 | 100% |
|  | HR+/HER2- | 0.70 | 0.50, 0.84 | 0.73 | 0.74 | 3.9E-2* | 0.32 | 0.44 | 74% |
|  | HR-/HER2- | 0.70 | 0.52, 0.83 | 0.78 | 0.63 | 2.0E-2* | 0.56 | 0.65 | 69% |

| **Metric** | **Subtype** | **AUC** | **CI** | **Sens** | **Spec** | ***p* value** | **Prec** | **F_mea_** | **Acc.** |
| --- | --- | --- | --- | --- | --- | --- | --- | --- | --- |
| ΔADC_02_ | HR+/HER2+ | 0.57 | 0.36, 0.82 | 0.33 | 1.0 | 5.4E-1 | 1.0 | 0.50 | 73% |
|  | HR-/HER2+ | 0.48 | 0.17, 0.75 | 1.0 | 0.45 | 9.8E-1 | 1.0 | 0.63 | 57% |
|  | HR+/HER2- | 0.58 | 0.33, 0.77 | 0.64 | 0.74 | 4.1E-1 | 0.29 | 0.40 | 72% |
|  | HR-/HER2- | 0.55 | 0.38, 0.74 | 0.67 | 0.5 | 5.6E-1 | 0.44 | 0.53 | 56% |
| ΔFTV_02_ | HR+/HER2+ | 0.47 | 0.26, 0.72 | 0.33 | 0.89 | 7.8E-1 | 0.67 | 0.44 | 67% |
|  | HR-/HER2+ | 0.88 | 0.58, 1.0 | 0.82 | 1.0 | 6.0E-2 | 1.0 | 0.90 | 86% |
|  | HR+/HER2- | 0.64 | 0.44, 0.79 | 0.55 | 0.72 | 1.5E-1 | 0.25 | 0.34 | 70% |
|  | HR-/HER2- | 0.65 | 0.46, 0.81 | 0.67 | 0.70 | 9.1E-2 | 0.57 | 0.62 | 67% |
